# A Cloth Facemask Causes No Major Respiratory or Cardiovascular Perturbations during Moderate to Heavy Exercise

**DOI:** 10.1101/2021.12.14.21267800

**Authors:** Natália Mendes Guardieiro, Gabriel Barreto, Felipe Miguel Marticorena, Tamires Nunes Oliveira, Luana Farias de Oliveira, Ana Lucia de Sá Pinto, Danilo Marcelo Leite do Prado, Bryan Saunders, Bruno Gualano

## Abstract

**Objectives:** To investigate whether wearing a cloth facemask could affect physiological and perceptual responses to exercise at distinct exercise intensities in non-trained men and women.

**Methods:** In a crossover design, participants (17 men and 18 women) underwent a progressive square-wave test at four intensities (i. at 80% of the ventilatory anerobic threshold [80%VAT]; ii. at VAT; iii. at the respiratory compensation point [RCP]; iv. at exercise peak [Peak] to exhaustion), with or without a triple-layered cloth mask (Mask or No-Mask). Several physiological, metabolic and perceptual measures were analyzed.

**Results:** Mask reduced inspiratory capacity at all exercise intensities vs. No-Mask (p<0.0001), irrespective of sex. Mask reduced respiratory frequency vs. No-Mask (p=0.001) at Peak (−8.3 breaths·min^-1^; CI: -5.8, -10.8), RCP (−6.9 breaths·min^-1^; CI: -4.6, -9.2) and VAT (−6.5 breaths·min^-1^; CI: -4.1, -8.8), but not at Baseline or at 80%VAT. Mask also reduced tidal volume (p<0.0001) at both RCP (−0.5L; CI: -0.3, -0.6) and Peak (−0.8L; CI: -0.6, -0.9), but not at Baseline, 80%VAT or VAT. Shallow breathing index was increased with Mask at Peak compared to No-Mask (11.3; CI: 7.5, 15.1), but not at any other intensities. Mask did not change heart rate, lactate, ratings of perceived exertion, blood pressure or oxygen saturation.

**Conclusions:** Wearing a cloth facemask during exercise at moderate to heavy intensities is unlikely to incur significant respiratory or cardiovascular changes, irrespective of sex. These data can inform new exercise recommendations for health during the COVID-19 pandemic and debunk unfounded allegations of harmful effects of masks during exercise. ClinicalTrials.gov: NCT04887714

**What are the new findings?:**

✓ Using a progressive square-wave test, we showed that wearing a cloth facemask during exercise increased breathing difficulty, but this was dependent upon the exercise intensity.
✓ Respiratory variables (e.g., inspiratory capacity, respiratory frequency, shallow breathing index) were affected at higher rather than lower intensities.
✓ Mask wearing did not change heart rate, lactate, ratings of perceived exertion, blood pressure or oxygen saturation at any exercise intensity.
✓ There were no substantial sex differences on the effects of mask wearing during exercise.

**How might it impact on clinical practice in the future?:**

✓ These data can debunk unfounded allegations on harmful effects of masks during exercise, and help inform new exercise recommendations for health during the COVID-19 pandemic, particularly where facemasks remain necessary.

## Introduction

The use of face masks is one of the most effective non-pharmacological strategy to prevent SARS-CoV-2 infections [1, 2]. The recent resurgence of cases and deaths worldwide has led to some decision-makers to re-issue mask orders to contain the disease, suggesting that this safety tool will remain important as long as the pandemic is not fully mitigated [3], and possibly after it is under control, as in several cultures employing facemasks as a routine practice to protect against several health threats before COVID-19 outbreak [4]. Wearing a facemask is recommended even during exercise, particularly at indoor fitness facilities and gyms, where COVID-19 outbreaks have been reported [5]. Nevertheless, the physiological impact of facemasks during exercise remains underexplored.

A facemask may reduce the ability to breathe comfortably during exercise, which has been confirmed by some [6, 7], but not all [8] studies. It is possible to conjecture that the effects of wearing a mask on cardiorespiratory responses may manifest during exhaustive high-intensity exercise, but not (or less) during low- to moderate-intensity exercise. In fact, Driver et al. [6] provide preliminary evidence that the effect of facemasks is dependent on exercise intensity, with differences in oxygen saturation, ratings of perceived exertion (RPE) and dyspnoea occurring at different stages of an incremental cardiopulmonary test as exercise intensity increased. However, short-stage maximal incremental tests do not normalize the physiological responses to exercise in relation to the gas exchange or blood acid-base profiles [9 10], since %VO_2max_ at the ventilatory thresholds largely varies between individuals [9], hampering accurate determination of exercise intensities and ultimately confounding data interpretation [10, 11]. To overcome this limitation, constant-load tests based on the dynamic behaviour of the pulmonary gas exchange and blood acid-base status have been recommended to accurately determine exercise intensity domains (i.e., moderate, heavy, severe, and extreme) [10]. This approach allows determination of whether facemasks affect physiological and perceptual parameters at different, well-defined exercise intensities, helping tailor exercise prescription for health that can minimize any negative effects of wearing a mask on cardiorespiratory responses.

Another remaining question is whether women and men respond differently to mask wearing during exercise. Generally, women have smaller lungs and airways, which limits their ability to generate expiratory flow [12, 13], which results in reduced ventilation during exercise compared to men. Women also have lower oxygen (O_2_) carrying capacity, maximum cardiac output, and arteriovenous O_2_ difference [14]. Considering the number of physiological and morphological sex differences to exercise, one could speculate that any physiological perturbations brought about wearing a mask during exercise could be greater in women, since men have an overall higher cardiorespiratory reserve.

This study aimed to investigate whether wearing a cloth facemask could affect physiological and perceptual responses to exercise at distinct exercise intensities in non-trained individuals. A secondary aim was to test whether sex differences influenced these responses.

## Methods

### Ethics statement

The protocol was approved by the institutional ethics committee. Written informed consent was obtained before participants’ enrollment.

### Study design and setting

This was a crossover study (clinicaltrials.gov: NCT04887714) performed at an intrahospital, exercise physiology laboratory in São Paulo, Brazil. Data collection took place between April and November 2021.

### Participants

Men and women not engaged in competitive sports (*i*.*e*., non-trained) were eligible for this study. Exclusion criteria included any cardiac, pulmonary, and rheumatologic diseases, musculoskeletal limitations, and BMI <18.5 or >30 kg·m^2^. A total of 18 men and 20 women entered the study, although 3 dropped out for reasons unrelated to the study. Thirty-five individuals (17 men and 18 women) completed all main sessions (age: women: 28±5 y, men: 30±4 y; BMI: women: 22.9±2.0 kg·m^2^, men: 24.5±2.6 kg·m^2^) and were analyzed. According to the short form of the International Physical Activity Questionnaire (IPAQ) instrument [15], 31 participants were physically active, whereas 4 were inactive.

### Experimental design

Participants attended the laboratory on three separate occasions, separated by a minimum of 48 h, at the same time of day to account for circadian variation [16]. The first visit consisted of an incremental cardiopulmonary running test to exhaustion to determine peak oxygen uptake (V□O_2peak_) and ventilatory thresholds. The remaining two main visits consisted of a running progressive square-wave test (PSWT), performed with or without the use of a triple-layered cloth facemask (Fashion Masks, São Paulo, Brazil). This facemask was chosen because it is widely accessible, recommended to the general public by the WHO and appropriate for exercise (https://www.cdc.gov/coronavirus/2019-ncov/prevent-getting-sick/cloth-face-cover-guidance.html). The outer layer was waterproof polyester fabric, the middle layer a polypropylene filter, and the inner layer was absorbable cotton. The size was “one size fits all” and was thus identical for all participants. Athletes were required to keep the mask firmly in place over the nose, mouth, and chin during the entire session. The order of sessions was determined by an individual not involved in data collection. Blocks of two individuals were allocated to the two possible orders (Mask–No Mask; No Mask–Mask) using a random number generator (https://www.randomizer.org/) to ensure the study was counterbalanced. Since the investigators could see that the participants were wearing a cloth facemask or not, the session order was provided directly to the research team. All participants were habituated to wear a mask during their daily routines due to mandates, but not specifically during exercise. Participants were requested to refrain from strenuous exercise, caffeine and alcohol, and replicated their diet, in the 24 h prior to each visit.

### Cardiorespiratory Exercise Test

Immediately prior to the cardiorespiratory exercise test, participants performed a pulmonary function test according to recommendations [17]. The cardiorespiratory exercise test was performed on a motorized treadmill (Centurion 300, Micromed, Brazil). For men, the test started at 5 km·h^-1^ and increased speed (1 km·h^-1^·min^-1^) up to a maximum velocity of 14 km·h^-1^. For women, the test started at 4 km·h^-1^ and increased speed (1 km·h^-1^·min^-1^) up to 13 km·h^-1^. For those participants who reached these maximal speeds, there was a subsequent increase in inclination (2%·min^-1^) until exhaustion. Ventilatory and gas exchange measurements were recorded continuously throughout the test using a breath-by-breath system (MetaLyzer 3B, Cortex, Germany), as was heart rate (HR; ergo PC elite, Micromed, Brazil). Maximal effort was determined according to published criteria [18] and individual V□O_2peak_ was determined as the V□O_2_ averaged over the final 30 s.

### Progressive square-wave test (PSWT)

Data from the cardiorespiratory exercise test was used to determine exercise workload for the square wave treadmill test according to the ventilatory anaerobic threshold (VAT) and the respiratory compensation point (RCP). All thresholds were determined by the same respiratory physiologist with clinical experience in the area. The PSWT protocol was performed on a motorized treadmill (Centurion, model 200, Micromed, Brazil) and consisted of three 5-min stages at workloads equivalent to 1) 80%VAT, 2) VAT, and 3) RCP. These stages represented moderate, heavy and severe domains [10] and corresponded to 41±9%, 53±9% and 81±8% of V□O_2peak_ of the volunteers. Participants then completed a final stage to exhaustion at a running speed equivalent to the maximum achieved during the cardiorespiratory exercise test (Peak). Ventilatory and gas exchange measurements were recorded continuously throughout, with the spirometer mask placed over the cloth facemask.

To determine the effect of the mask on pattern of change in operating lung volumes we evaluated end-expiratory volume to functional vital capacity ratio (EELV/FVC). Inspiratory capacity was determined at rest and at the end of each exercise stage during the PSWT. Ventilatory constraint was evaluated as the difference between inspiratory capacity at rest and at each exercise workload [19]. Ventilatory efficiency was determined using the ventilatory equivalent for carbon dioxide (V□_E_/V□CO_2_) and end-tidal carbo dioxide pressure (PetCO_2_) during each stage. Breathing pattern was evaluated during each stage using the breathing frequency to tidal volume ratio (BF/TV) ratio [20].

RPE were assessed at the end of each stage with participants pointing to a chart using the 6- to 20-point Borg scale [21]. Heart rate was monitored continuously throughout (ergo PC elite, Micromed, Brazil). A fingertip blood sample (20 μL) was collected at baseline, at the end of each stage and 4-min post-exhaustion for the subsequent analysis of lactate. Blood was homogenized in the same volume of 2% NaF, centrifuged at 2000 g for 5 min before plasma was removed and stored at -20^°^C until analysis. Plasma lactate was determined spectrophotometrically using an enzymatic-colorimetric method (Katal, Interteck, Brazil).

### Subjective Perception of Discomfort Questionnaire

Participants completed a questionnaire [22] following the completion of the PSWT to rate their perception of ten sensations of discomfort related to the facemask: humidity, heat, breathing resistance, itchiness, tightness, saltiness, feeling unfit, odor and fatigue. They were also required to rate their overall feeling of discomfort related to the facemask on a scale from 0 to 10, with 0–3 representing “Comfortable”, >3–7 representing “Uncomfortable” and >7 representing “Extremely uncomfortable”.

### Statistical analyses

Lactate data from 2 individuals (1 male, 1 female) who did not complete the third stage were excluded from the RCP and Peak analyses. Furthermore, 1 male and 1 female reported extreme discomfort with the PSWT mask and stopped exercising before volitional exhaustion; and were excluded from the Peak analysis for all outcomes. The same 4 individuals were excluded from the TTE analysis. Lactate data were log10 transformed before mixed model analysis, turning the model into an exponential data mixed model, and back transformed through exponentiation for the final reporting of data. Whenever outlying data points were considered improbable (e.g., a value of 50 mmHg for systolic blood pressure), they were considered measurement or transcription error and were excluded.

Repeated measures Mixed Model ANOVAs were performed with condition (Mask, No-Mask), sex and exercise intensity (Baseline [except RPE], 80%VAT, VAT, RCP, Peak) as fixed factors and individuals as random factors. Exceptions were Spirometry, TTE and Questionnaire related outcomes, which were not repeated measures and, therefore, timing was not included as a fixed factor. For TTE, the model was corrected by treatment order, since participants were not familiarized to the protocol. When a significant main effect or interaction was detected (accepted at p≤0.05), post-hoc pairwise comparisons were performed with Tukey’s adjustment. All analyses were performed with the RStudio software (Rstudio 1.4.11003, PBC, MA) using the “lmer” function of the lmerTest package and “emmeans” function of the emmeans package. Data are expressed as estimated differences and 95% confidence intervals (CI), and data in figures are represented as mean±1standard deviation.

### Patient and Public Involvement statement

Participants were not involved in the design or in performing the study.

## Results

### Cardiorespiratory Exercise Test

Participants’ characteristics and respiratory data at the ventilatory thresholds and calculated stages are presented in Table 1.

**Table 1.**
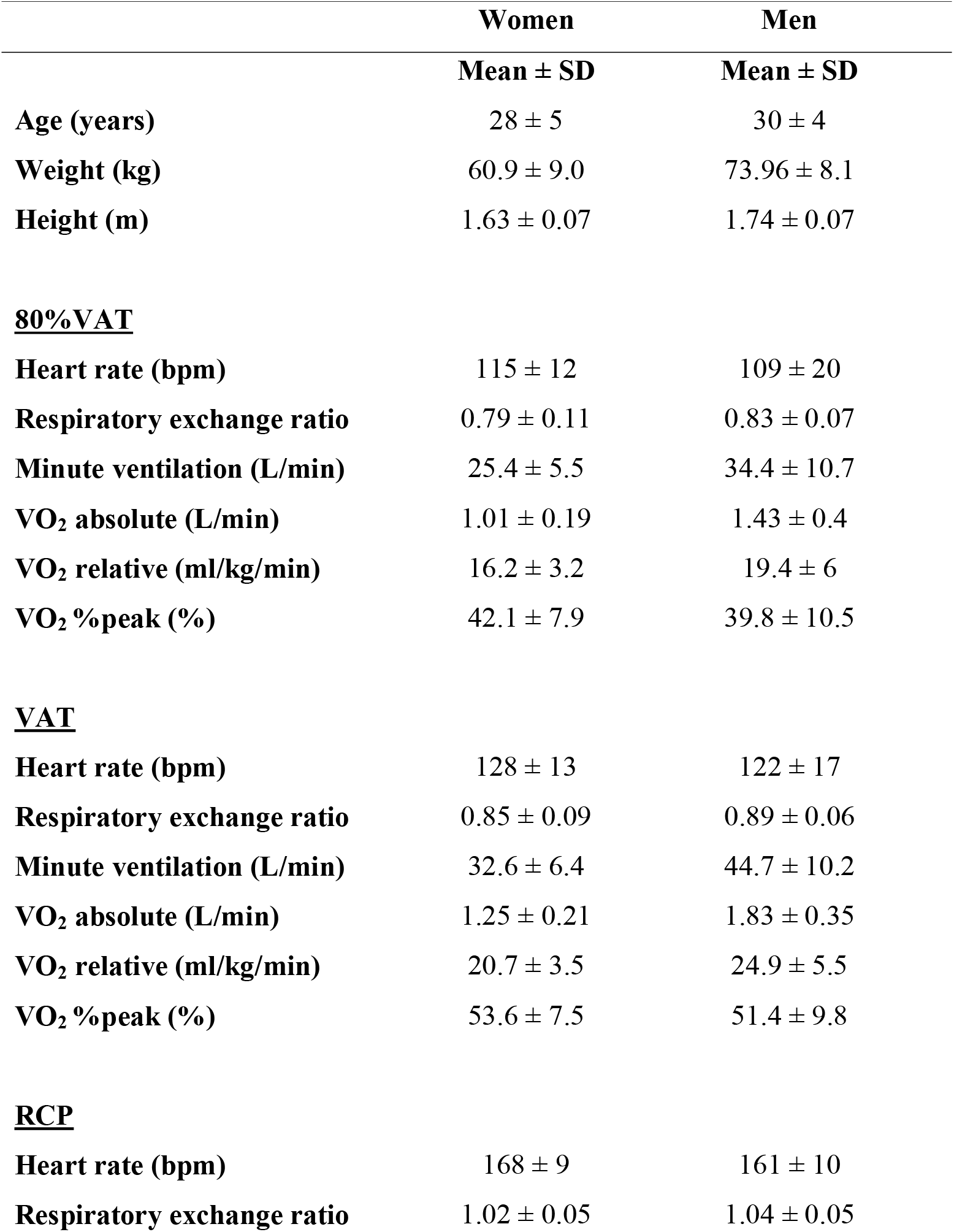
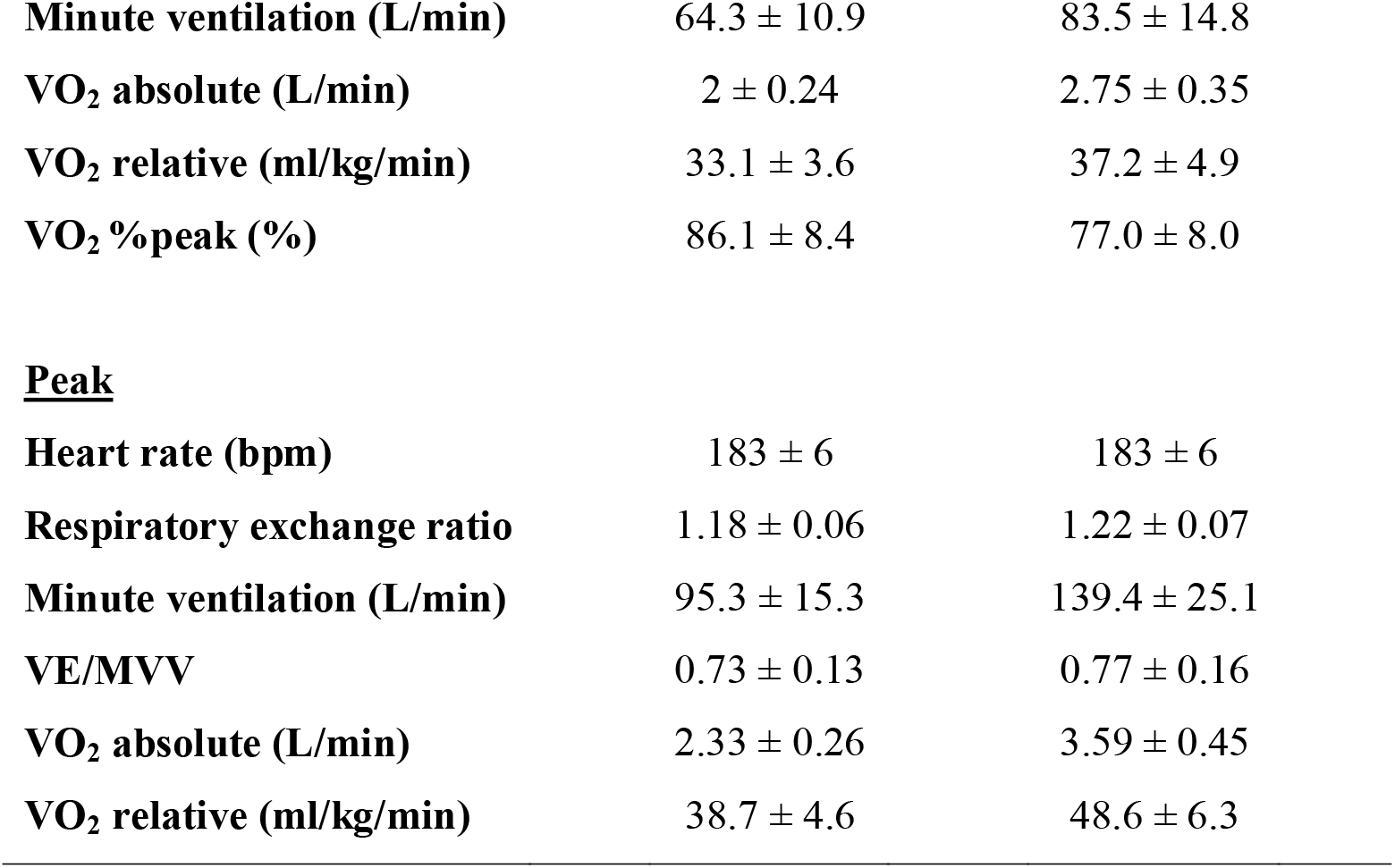
Participants’ characteristics and cardiorespiratory data from the cardiopulmonary exercise test. VE/MVV: minute ventilation/maximum voluntary ventilation ratio.

|  | **Women** | **Men** |
| --- | --- | --- |
|  | **Mean ± SD** | **Mean ± SD** |
| **Age (years)** | 28 ± 5 | 30 ± 4 |
| **Weight (kg)** | 60.9 ± 9.0 | 73.96 ± 8.1 |
| **Height (m)** | 1.63 ± 0.07 | 1.74 ± 0.07 |
| **<u>80% VAT</u>** |  |  |
| **Heart rate (bpm)** | 115 ± 12 | 109 ± 20 |
| **Respiratory exchange ratio** | 0.79 ± 0.11 | 0.83 ± 0.07 |
| **Minute ventilation (L/min)** | 25.4 ± 5.5 | 34.4 ± 10.7 |
| **VO<sub>2</sub> absolute (L/min)** | 1.01 ± 0.19 | 1.43 ± 0.4 |
| **VO<sub>2</sub> relative (ml/kg/min)** | 16.2 ± 3.2 | 19.4 ± 6 |
| **VO<sub>2</sub> %peak (%)** | 42.1 ± 7.9 | 39.8 ± 10.5 |
| **<u>VAT</u>** |  |  |
| **Heart rate (bpm)** | 128 ± 13 | 122 ± 17 |
| **Respiratory exchange ratio** | 0.85 ± 0.09 | 0.89 ± 0.06 |
| **Minute ventilation (L/min)** | 32.6 ± 6.4 | 44.7 ± 10.2 |
| **VO<sub>2</sub> absolute (L/min)** | 1.25 ± 0.21 | 1.83 ± 0.35 |
| **VO<sub>2</sub> relative (ml/kg/min)** | 20.7 ± 3.5 | 24.9 ± 5.5 |
| **VO<sub>2</sub> %peak (%)** | 53.6 ± 7.5 | 51.4 ± 9.8 |
| **<u>RCP</u>** |  |  |
| **Heart rate (bpm)** | 168 ± 9 | 161 ± 10 |
| **Respiratory exchange ratio** | 1.02 ± 0.05 | 1.04 ± 0.05 |
| **Minute ventilation (L/min)** | 64.3 ± 10.9 | 83.5 ± 14.8 |
| **VO<sub>2</sub> absolute (L/min)** | 2 ± 0.24 | 2.75 ± 0.35 |
| **VO<sub>2</sub> relative (ml/kg/min)** | 33.1 ± 3.6 | 37.2 ± 4.9 |
| **VO<sub>2</sub> %peak (%)** | 86.1 ± 8.4 | 77.0 ± 8.0 |
| **Peak** |  |  |
| **Heart rate (bpm)** | 183 ± 6 | 183 ± 6 |
| **Respiratory exchange ratio** | 1.18 ± 0.06 | 1.22 ± 0.07 |
| **Minute ventilation (L/min)** | 95.3 ± 15.3 | 139.4 ± 25.1 |
| **VE/MVV** | 0.73 ± 0.13 | 0.77 ± 0.16 |
| **VO<sub>2</sub> absolute (L/min)** | 2.33 ± 0.26 | 3.59 ± 0.45 |
| **VO<sub>2</sub> relative (ml/kg/min)** | 38.7 ± 4.6 | 48.6 ± 6.3 |

### Progressive square-wave test (PSWT)

#### Inspiratory capacity (IC)

Mask reduced IC at all exercise intensities (interaction effect condition*timing: F=8.6, p<0.0001, Figure 1, Panel A) compared to No-Mask irrespective of sex (80%VAT: -0.4 L; CI: -0.2, -0.6; VAT: -0.5 L; CI: -0.4, -0.7; RCP: -0.7 L; CI: -0.5, -0.9; Peak: -1.0 L; CI: -0.8, - 1.2), except at Baseline (−0.2 L; CI: 0.0, -0.4).

**Figure 1.**
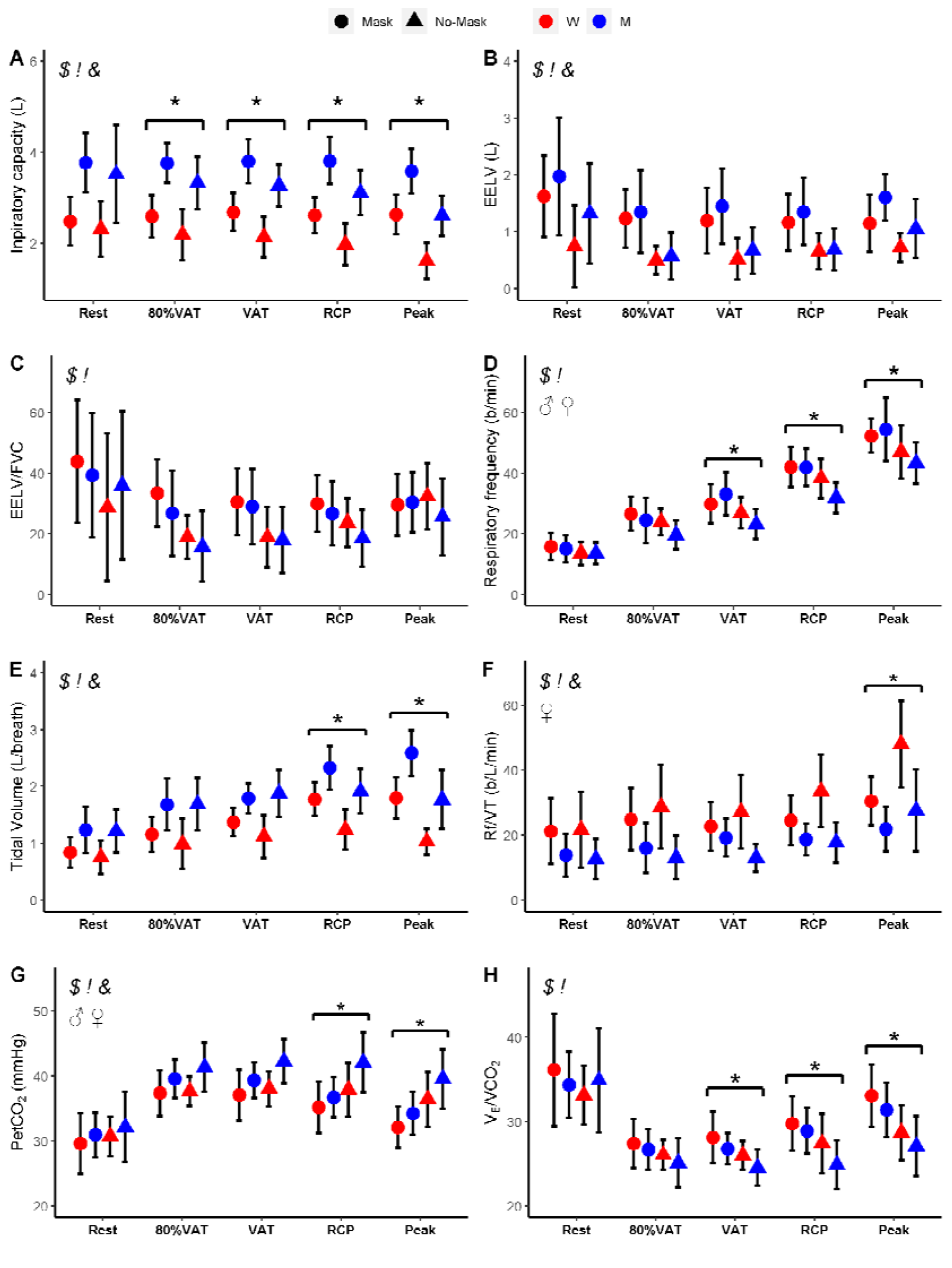
Inspiratory capacity, end expiratory lung volume (EELV), EELV by forced vital capacity ratio (EELV/FVC), respiratory frequency, tidal volume, respiratory frequency by tidal volume, CO_2_ partial pressure (PetCO_2_) and ventilatory equivalent for carbon dioxide (V_E_/VCO_2)_ data expressed as mean ± 1 standard deviation. ^$^ main effect of Mask; ^!^ main effect of intensity; ^&^ main effect of sex; ^*^ condition*intensity interaction; ^♂^ condition*intensity interaction for men; ^♀^ condition*intensity interaction for women.

#### End expiratory lung volume (EELV) and EELV/Forced Vital capacity (EELV/FVC)

Mask did not influence EELV or EELV/FVC, irrespective of exercise intensities and sex (all p≥0.1, Figure 1, Panel B and C).

#### Respiratory Frequency (Rf)

Mask reduced Rf vs. No-Mask (interaction effect of condition*timing: F=4.6, p=0.001, Figure 1, Panel D) at Peak (−8.3 breaths·min^-1^; CI: -5.8, -10.8), RCP (−6.9 breaths·min^-1^; CI: - 4.6, -9.2) and VAT (−6.5 breaths·min^-1^; CI: -4.1, -8.8), but not at Baseline or at 80%VAT (both p≥0.06). Rf was reduced similarly in men (−7.5 breaths·min^-1^; CI: -6.0, -9.0) and women (−3.4 breaths·min^-1^; CI: -1.9, -4.9) with Mask (Figure 1, Panel D).

#### Tidal Volume (VT)

Mask reduced VT (interaction effect of condition*intensity, F=18.3, p<0.0001, Figure 1, Panel E) at both RCP (−0.5L; CI: -0.3, -0.6) and Peak (−0.8L; CI: -0.6, -0.9), but not at Baseline, 80%VAT or VAT (all p≥0.97). Sex had no influence on the effects of Mask on VT (p=0.053).

#### Tobin index (Rf/VT or shallow breathing index)

Mask increased the Tobin index at Peak compared to No-Mask (+11.3; CI: 7.5, 15.1), but not at any other intensity (all p≥0.4, interaction effect of condition*intensity: F=7.3, p<0.0001). Rf/VT in men was not affected by Mask, whereas it was increased in Mask vs. No-Mask for women (interaction effect of condition*sex: F=25.1, p<0.0001; men: +1.1; CI: -1.1, 3.3; women: +6.9; CI: 4.7, 9.1; Figure 1, Panel F).

#### *PetCO*_*2*_

Mask increased PetCO_2_ at both RCP (+4.0 mmHg; CI: 2.8, 5.3, Figure 1, Panel G) and Peak (+4.9 mmHg; CI: 3.5, 6.3) compared to No-Mask (interaction effect condition*intensity: F = 6.8, p<0.0001), but had no effect at Baseline, 80%VAT or VAT (all p≥0.09). The effect of Mask on PetCO_2_ increases was comparable in men (+3.4 mmHg; CI: 2.5, 4.2) and women (+1.9 mmHg; CI: 1.1, 2.7).

#### V□_E_/ V□ CO_2_

Mask reduced VE/VCO_2_ at the three highest intensities compared to No-Mask (interaction effect of condition*intensity, F=3.7, p=0.006): VAT (−2.2; CI: -0.9, -3.5), RCP (−3.2; CI: -1.9, -4.5), and Peak (−4.4; CI: -3.0, -5.8) (Figure 1, Panel H), irrespective of sex.

#### Heart rate during exercise (HR)

No effects of Mask at any exercise intensity were seen for HR. Despite an interaction effect of condition*sex for HR (F=4.6, p=0.03), post-hoc comparisons did not show any significant differences (all p≥0.4). (Figure 2, Panel A).

**Figure 2.**
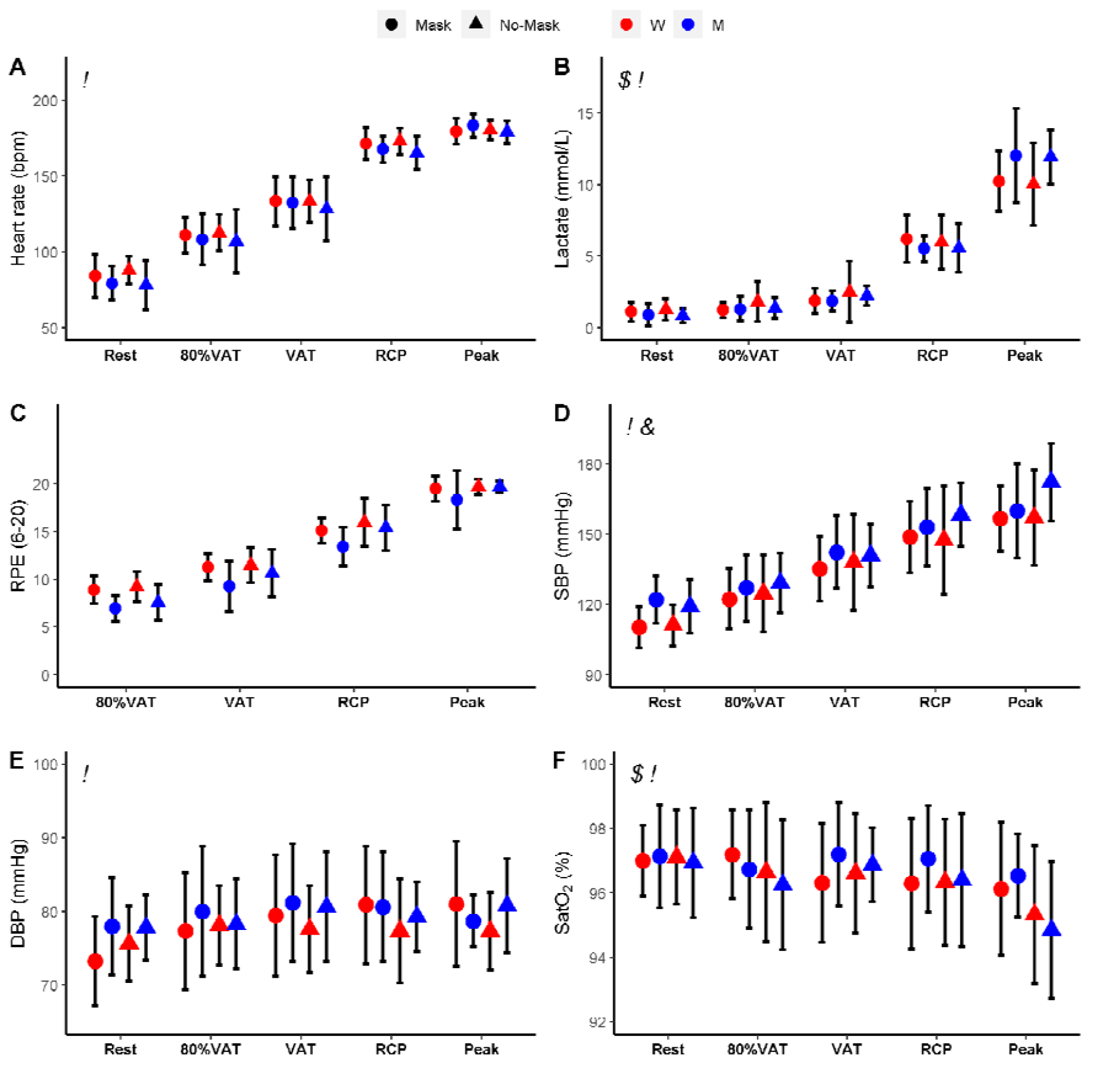
Heart rate, lactate, rating of perceived effort (RPE), systolic (SBP) and diastolic (DBP), and oxygen saturation (SatO_2_) expressed as mean ± 1 standard deviation. ^$^ main effect of Mask; ^!^ main effect of intensity; ^&^ main effect of sex; ^♂^ condition*intensity interaction for men.

#### Lactate

Mask did not affect lactate measures at any exercise intensities, irrespective of sex (Figure 2, Panel B).

#### Ratings of Perceived Exertion (RPE)

Mask did not influence RPE at any exercise intensity. However, Mask increased RPE for men, but not women (interaction effect of condition*sex, F=6.2, p=0.01, men: +1.4; CI: 0.9, 2.0; women: +0.4; CI: -0.1, 1.0; Figure 2, Panel C).

#### Blood pressure

Mask did not affect both systolic or diastolic blood pressure at any exercise intensity (Figure 2, Panel D and E), regardless of sex.

#### Oxygen saturation (SatO_2_)

Mask did not affect SatO_2_ at any exercise intensity (Figure 2, Panel F), regardless of sex.

#### Time-to-exhaustion (TTE)

Mask reduced TTE compared to No-Mask (−34.5 s; CI: -17.0, -52.1; main effect of condition: F=14.9, p=0.0007, Figure 3), with no condition*sex interaction.

**Figure 3.**
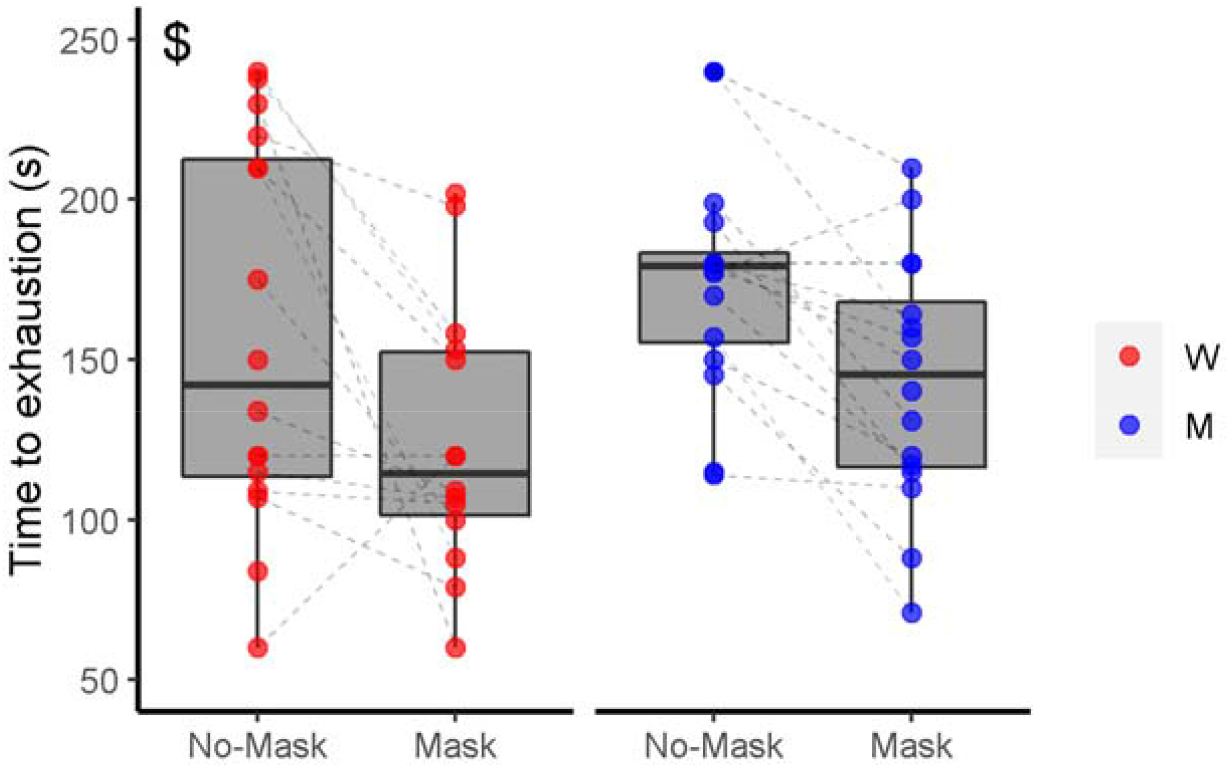
Time-to-exhaustion during the final stage. Dashed lines connect individual performance data between No-Mask and Mask condition. ^$^ main effect of Mask

#### Forced Vital Capacity at rest (FVC)

Mask reduced FVC compared to No-Mask (−1.8L; CI: -1.1, -1.5; condition: F=117.7, p<0.0001), with no condition*sex interaction (Table 2).

**Table 2.** Spirometry at rest and subjective questionnaire outcomes.

|  | **Women** |  | **Men** |  |
| --- | --- | --- | --- | --- |
|  | **No-Mask** | **Mask** | **No-Mask** | **Mask** |
|  | **Mean ± SD** | **Mean ± SD** | **Mean ± SD** | **Mean ± SD** |
| **<u>Spirometry</u>** |  |  |  |  |
| **FEV1 (L)<sup>\$!</sup>** | 3.4 ± 0.4 | 2.3 ± 0.5 | 4.5 ± 0.5 | 3.2 ± 0.6 |
| **FVC (L)<sup>\$!</sup>** | 3.8 ± 0.6 | 2.6 ± 0.6 | 5 ± 0.7 | 3.7 ± 0.7 |
| **FVC/FEV1(%)** | 90.7 ± 7.7 | 90.5 ± 7.2 | 90.8 ± 6.7 | 87.3 ± 8.4 |
| **PEF (L/s)<sup>\$!</sup>** | 7.7 ± 1.8 | 4.4 ± 1.1 | 9.2 ± 1.9 | 5.7 ± 1.6 |
| **<u>Questionnaire</u>** |  |  |  |  |
| **Tightness** | 3.4 ± 2.4 | 4.2 ± 3.4 | 2.5 ± 2.2 | 3.4 ± 2.5 |
| **Heat<sup>\$</sup>** | 3.6 ± 2.1 | 6.2 ± 2.5 | 3.3 ± 2.2 | 5.9 ± 2.3 |
| **Itchiness** | 0.8 ± 1.1 | 1.1 ± 2.2 | 1.6 ± 2.7 | 2.8 ± 3.1 |
| **Misfitting<sup>\$</sup>** | 1.8 ± 2.3 | 2.6 ± 2.5 | 1.6 ± 2.1 | 4.1 ± 2.7 |
| **Discomfort<sup>\$</sup>** | 4.5 ± 1.4 | 7.9 ± 2.3 | 4.2 ± 2.9 | 7.1 ± 2.6 |
| **Fatigue<sup>\$</sup>** | 5.2 ± 3.1 | 8.2 ± 1.8 | 5.0 ± 2.6 | 8.6 ± 2.4 |
| **Odour** | 1.2 ± 2.4 | 0.9 ± 1.8 | 0.9 ± 1.3 | 1.7 ± 2.4 |
| **Resistance<sup>\$</sup>** | 3.4 ± 2.3 | 8.6 ± 1.2 | 4.0 ± 2.8 | 8.4 ± 1.3 |
| **Saltiness<sup>\$!</sup>** | 0.3 ± 0.5 | 0.7 ± 1.6 | 1.0 ± 1.8 | 2.7 ± 3.5 |
| **Humidity<sup>\$</sup>** | 3.7 ± 2.6 | 5.3 ± 2.3 | 4.1 ± 1.9 | 6.6 ± 2.4 |

FEV1, forced expiratory volume in 1 second; FVC, forced vital capacity; FVCbyFEV1,
forced vital capacity by FEV1 ratio; PEF, peak expiratory flow.
Significance labels: <sup>\$</sup>, main effect of condition; <sup>!</sup>, main effect of sex; <sup>&</sup>, interaction effect

#### Forced expiratory volume in 1 second at rest (FEV1)

Mask reduced FEV1 compared to No-Mask (−1.2; CI: -1.0, -1.4; main effect of condition: F=156.2, p<0.0001), with no condition*sex interaction (Table 2).

#### FVC/FEV1 at rest (FVC by FEV1 ratio)

Mask did not influence the FVC/FEV1 ratio, irrespective of sex (Table 2).

#### Peak expiratory flow at rest (PEF)

Mask reduced PEF compared to No-Mask (−3.4; CI: -2.8, -4.0, main effect of condition: F=122.1, p<0.0001), independently of sex (Table 2).

#### Subjective perception questionnaire

Mask increased the subjective feelings of Heat, Misfitting, Discomfort, Fatigue, Resistance, Saltiness and Humidity (all p≤0.01), but did not affect feelings of Saltiness, Tightness or Itchiness (all p≥0.1). No interaction between condition and sex was detected (Table 2).

## Discussion

This study showed that breathing difficulty with a cloth facemask is dependent upon the exercise intensity, with lower distress at less severe intensities. Mask wearing did not substantially affect physiological or metabolic variables during exercise, regardless of sex and intensity. From a practical perspective, these data suggest that use of a cloth facemask for protecting individuals from SARS-CoV-2 infections should not be a barrier to the engagement in moderate-to-heavy physical activity.

The main novelty of the current study is that we assessed the influence of wearing a mask on respiratory and cardiovascular variables across several exercise intensities, spanning moderate to severe domains [10]. Wearing a mask did not modify most respiratory variables in the moderate to heavy exercise domains, with only inspiratory capacity reduced at 80%VAT with the mask. Our findings suggest that reductions in inspiratory capacity early in exercise are reflective of a decrease in contractile power of the inspiratory muscles. This inspiratory distress may place a greater strain on the respiratory muscles to maintain breathing requirements during exercise. This was not seen in the moderate to heavy domains but was manifested in an inability to maintain the physiological increases in respiratory frequency and tidal volume at the higher intensities. For example, during VAT, although breathing frequency was reduced, this was likely compensated by increases in tidal volume as evidenced by no differences between conditions, suggesting that the cloth facemask did not negatively inhibit the ability of the respiratory system to work at moderate, heavy and severe exercise intensities. Indeed, this might explain the reduced V□_E_/V□CO_2_ with a mask. At higher intensities, however, both respiratory frequency and tidal volume were reduced with a mask, which may have led to an inability to maintain respiratory homeostasis and affected subsequent performance. Our data are in agreement with previous studies showing a reduced exercise capacity with different facemasks [6 22], though others have shown no negative impact [8, 23]. It is possible that different types of facemasks and respirators (e.g., FFP2/N95, surgical, cloth facemasks) and participants’ fitness level may have contributed to these conflicting findings.

Despite some changes in respiratory variables, no cardiovascular measure was affected using a cloth facemask. Even during the highest exercise intensity domains, there were no changes in heart rate, or systolic and diastolic blood pressure when wearing a cloth facemask. Moreover, the slight overall reduction in blood oxygen saturation (not detected at individual exercise intensities) was not clinically meaningful, supporting previous studies showing no effect of wearing a facemask on oxygen saturation [24, 25]. These findings collectively can counteract misinformation during the COVID-19 pandemic, particularly relating to the use of masks during exercise and its supposed negative effects on cardiac overload, acid-base balance, and oxygen saturation [28].

We had speculated that any potential physiological effects associated with wearing a mask during exercise could be greater in women, who have an overall lower higher cardiorespiratory reserve than men, owing to classically described morphological and physiological sex-differences (e.g., smaller lungs, lower O_2_ carrying capacity and maximum cardiac output). This hypothesis was not confirmed, as the effects of the cloth facemask on physiological measures were mostly similar between women and men irrespective of exercise intensities. Exercise capacity was reduced by 23.9% in women and 17.8% in men when using a cloth facemask, with no differences between sexes. This suggests that the stress imposed by wearing a mask does not constitute a greater physiological or metabolic burden to women vs. men, despite the well-known sex-differences during exercise.

This study has several limitations. Although the measurement of respiratory variables during the PSWT provide novel information regarding the respiratory response during different intensities, participants were required to wear a facemask for breath-by-breath measures over the cloth facemask. This may have increased the discomfort felt by the participants and may also have led to some inaccuracies in measurements due to air escaping. The current data cannot be directly extrapolated to trained individuals; however, we felt it important to investigate this matter among a non-trained population, as there has been intense debate on the physiological repercussions and potential adverse effects of face masks in non-trained individuals. Since sufficient levels of physical activity prevent morbidities and mortality [29-31] and improve vaccine immunogenicity [32], it is important that mask mandates do not lead to a reduction in physical activity. In this regard, the present data provide relevant information that wearing a cloth facemask will not have a negative impact during exercise at moderate-to-heavy intensities, which is associated with a plethora of health-related benefits [33, 34]. Whether the negative perpetual feelings related to the use of masks may result in less adherence to exercise remains to be examined. Furthermore, the influence of mask wearing during exercise in clinical populations warrants investigation.

In conclusion, although wearing a cloth facemask may reduce maximal exercise capacity, it did not impose significant respiratory or cardiovascular changes during exercise performed at moderate-to-heavy intensities. These data have important practical implications as they can debunk unfounded allegations of harmful effects of facemasks during exercise and help inform new exercise recommendations for health during the COVID-19 pandemic.

## Data Availability

All data produced in the present study are available upon reasonable request to the authors

## Acknowledgments

The authors would like to thank all of the athletes for taking part in this research. The authors received no specific funding for this work. B.G (2017/13552-2), G.B. (2020/12036-3), T.N.O. (2020/04368-6) and B.S. (2016/50438-0) have been financially supported by Fundação de Amparo à Pesquisa do Estado de São Paulo. B.S. has also received a grant from Faculdade de Medicina da Universidade de São Paulo (2020.1.362.5.2).

## Notes

### Competing Interest Statement

The authors have declared no competing interest.

### Clinical Trial

NCT04887714

### Author Declarations

The protocol was approved by the ethics committee of the Hospital das Clinicas of the Faculty of Medicine of the University of Sao Paulo under the protocol 38569420.0.0000.0068

### Summary of Updates

Table 2 was revised. Also, we reduced the text in ~600 words.

